# Thalamic sonication in chronic disorders of consciousness: a mechanistic single-arm clinical trial

**DOI:** 10.64898/2026.05.26.26354167

**Authors:** Martin M. Monti, Amber R. Hopkins, Norman M. Spivak, Joshua A. Cain, Jeannette Gumarang, David Patterson, Emily R. Rosario, Caroline Schnakers

**Affiliations:** Department of Psychology, University of California Los Angeles, Los Angeles, CA, USA; Brain Injury Research Center (BIRC), Department of Neurosurgery, University of California Los Angeles, Los Angeles, CA, USA; UCLA-Caltech Medical Scientist Training Program, David Geffen School of Medicine, University of California Los Angeles, Los Angeles, CA, USA; Casa Colina Research Institute, Casa Colina Hospitals and Centers for Healthcare, Pomona, CA, USA; Physical Medicine & Rehabilitation, Casa Colina Hospitals and Centers for Healthcare, Pomona, CA, USA

**Keywords:** Disorders of Consciousness (DOC), transcranial Focused Ultrasound (tFUS), thalamus, case series

## Abstract

**Background:** Thalamic low-intensity transcranial focused ultrasound (tFUS) has shown promise for increasing behavioral responsiveness in disorders of consciousness (DOC), but no study has examined whether it can causally modulate the well-validated behavioral, electrophysiological, and metabolic biomarkers of DOC impairment.

**Methods:** Sixteen adult patients (44% Female; Age, M=37.81, SD=15.97) with a chronic DOC (Time Since Injury, M=3.39, SD=1.94 years) secondary to severe brain injury (TBI 44%, non-TBI 56%) underwent a 10-day inpatient, longitudinal, single-arm, open-label protocol. tFUS was delivered in a single session targeting the left central thalamus. Well-known behavioral (CRS-R), electrophysiological (EEG δ/β ratio), metabolic (18F-FDG PET), and polysomnographic outcomes were assessed at baseline and after sonication.

**Results:** The maximum CRS-R total score increased significantly following tFUS compared to baseline (M=13.27 vs. M=10.33; t(14)=7.407, p<0.001, d=1.913), as did the global EEG δ/β ratio (N=14; W=17, p=0.025, r=0.68), with the degree of frontal slowing positively predicting behavioral gains (τ_b_=0.51, p=0.016). Glucose metabolism decreased bilaterally in thalamus and frontal, temporal, and parietal cortices at both post-tFUS timepoints compared to baseline. Finally, N2 sleep increased by 33% following tFUS (N=11; t(10)=2.386, p=0.038, d=0.72), though this did not survive correction. No severe adverse events were observed.

**Conclusion:** Thalamic tFUS can causally modulate well-validated behavioral, electrophysiological, and metabolic biomarkers of DOC. The convergent inhibitory signature across these measures suggests a thalamocortical reset mechanism, complementing existing excitatory neuromodulation approaches and providing the mechanistic foundation for a large, randomized sham-controlled trial.

---

Despite an exciting landscape of interventional science,^1–3^ once patients with a Disorder of Consciousness (DOC)^4–6^ such as the Vegetative State (VS)^7^ and the Minimally Conscious State (MCS),^8^ stabilize in a chronic DOC, there are few therapeutic options available.^2,9^ Pharmacological interventions such as amantadine have shown some of the strongest results for use in prolonged DOC,^10^ but there are no comparable data for chronic patients. Zolpidem has been shown to be transiently (and paradoxically) awaken DOC patients in post-acute, mixed, and chronic cohorts, but only in ∼5% of cases.^11,12^ Thalamic Deep Brain Stimulation (DBS) has been used in acute and chronic DOC cohorts with varied, albeit at times striking, post-stimulation recovery.^13–16^ Nonetheless, eligibility rates in DOC have ranged from 45% to less than 13% of screened patients, in prospective clinical trials.^14,16^ Finally, transcranial Direct Current Stimulation (tDCS) ^17,18^ has also been shown, in rigorous double-blind settings, to be effective at increasing responsiveness in DOC patients, albeit mainly in MCS patients with traumatic or vascular etiologies.^19^

Low-intensity, reversible transcranial focused ultrasound (tFUS), has recently emerged as a novel neuromodulatory modality uniquely capable of targeting deep neural tissues, including those believed to be at the heart of DOC,^20^ without requiring surgical interventions. While the exact cellular and molecular mechanisms of neuromodulatory tFUS are not completely understood,^21^ sonication is believed to lead to transmembrane ionic fluxes *via* sound-induced tissue compression, tension, and/or sheering, which, in turn, lead to changes in the membrane-resting potential and activation of ionotropic voltage-gated channels.^22,23^ Neuromodulatory effects of tFUS in brain tissues have been demonstrated *in vitro*,^24,25^ in animal models,^26–29^ and in human volunteers.^30–32^ Thalamic tFUS specifically has been shown, in a rodent model of anesthesia, to accelerate the recovery of behavioral responsiveness,^28^ paralleling recent DBS work in non-human primate models of anesthesia^33,34^ and suggesting the potential for restoring responsiveness in states of reduced consciousness. Building on this preclinical foundation, thalamic tFUS has indeed been associated with increased behavioral responsiveness in proof-of-concept studies in acute and chronic DOC patients.^35–37^

In light of the above data, and the excellent safety profile of low-intensity focused ultrasound,^38,39^ we report a longitudinal study assessing the degree to which thalamic tFUS can causally modulate well-validated biomarkers of impaired consciousness in chronic DOC patients. Specifically, we adopt a mechanistic, open-label, longitudinal trial design in a cohort of 16 chronic DOC patients undergoing a 10-day protocol featuring repeated sampling of established neurobehavioral (i.e., Coma Recovery Scale-Revised; CRS-R),^40^ electrophysiological (i.e., EEG),^41,42^ and metabolic (i.e., 18F-fluorodeoxyglucose Positron Emission Tomography; ^18^F-FDG PET)^43,44^ biomarkers of DOC before and after thalamic tFUS.

## Methods

### Study Design

This was a single-arm, open-label, study of a consecutive chronic DOC sample, conducted between 2021 and 2023 at a tertiary referral rehabilitation center. All procedures were approved by the IRB of Casa Colina Hospital and Centers for Rehabilitation. For each patient, their Legally Authorized Representative provided written consent following local, state, and federal policy. This work was registered on ClinicalTrials.gov (NCT04921683).

### Participants

Key inclusion criteria were 18 years of age or older, a diagnosis of VS or MCS as determined by CRS-R,^40^ with time since injury (TSI) compatible with a diagnosis of chronic DOC (i.e., at least 1 year after traumatic brain injury [TBI] or 3 months after non-TBI [NTBI], respectively). Key exclusion criteria were a history of neurological disorders prior to the brain injury that led to a DOC, dependence on a ventilator, unsuitability for undergoing MRI and/or PET-CT examinations (e.g., having already received an exam involving the use of ionizing radiation in the 12 months prior to enrollment), unsuitability to undergo tFUS sonication (e.g., presence of non-bone implant or absence of bone under the expected positioning of the device by the left temporal bone window), and concurrent participation in another clinical trial.

### Protocol

The study featured a 10-day protocol, as an in-patient admission, inclusive of a 4.5-day baseline observation period and a 5.5-day follow-up period. As summarized in the schedule of activities (see Supplementary Materials, Figure 1), participants underwent multiple neurobehavioral, electroencephalographic, metabolic, and polysomnography assessments, before and after tFUS. Prior to tFUS, a total of 5 neurobehavioral assessments were acquired, following international guidelines,^5,6^ with the CRS-R, and at least 5 additional assessments were acquired following tFUS (i.e., follow-up). Five EEG assessments were conducted, two during baseline observation and three following the intervention. Each assessment consisted of a 10-min period of resting-state EEG. While the trial was originally conceived (and registered) to also feature an auditory violation paradigm^45^ during EEG recordings, this was never performed due to concerns relating to patient fatigue, particularly on the day of tFUS administration, which already featured two neurobehavioral and two resting state EEG acquisitions administered “back-to-back” before and after sonication. Three neurometabolic assessments were conducted, using ^18^F-FDG PET,^46,47^ one during baseline and two following tFUS. Finally, polysomnography was acquired very night using the Dreem 2 headband.^48,49^

**Figure 1.**
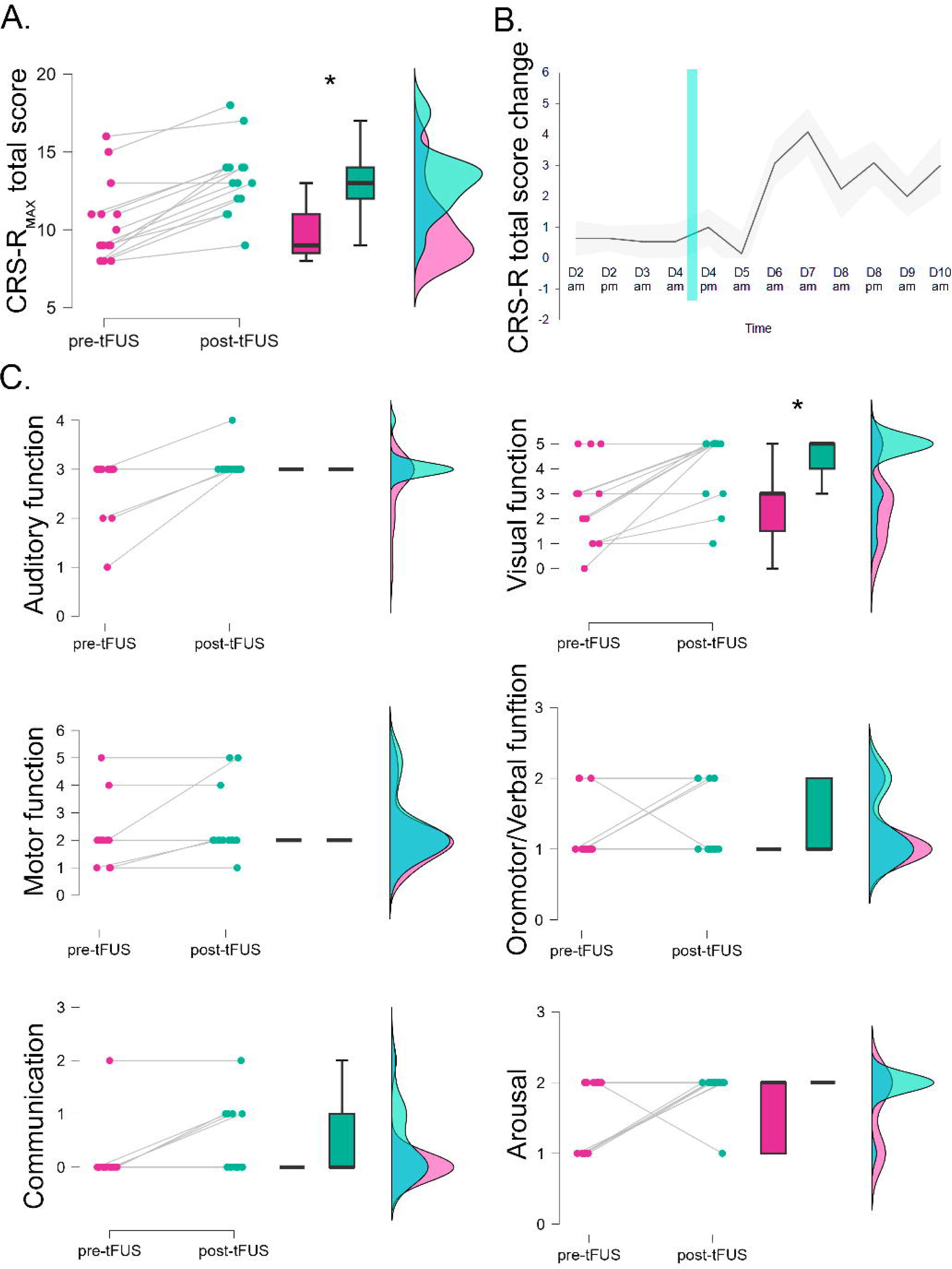
*Neurobehavioral results.* (A) Comparison of the maximum CRS-R total score following tFUS (post-tFUS) compared to the maximum CRS-R total score observed during the baseline (pre-tFUS). (B) Time-course, normalized to the first assessment of the average change in CRS-R total score at each assessment (the black line indicates the mean change, the gray shaded area represents the standard error band). (C) Same as quadrant A, for each CRS-R subscale separately.

### Study intervention

tFUS was delivered with a Brainsonix BX Pulsar 1002 device, operating at a fundamental frequency of 650 kHz, with a 61 mm curvature diameter piezoelectric element, a radial focus of approximately 0.25 cm and a longitudinal focal spread of ∼1.25 cm.^50^ To accommodate patient-specific neuroanatomy (e.g., degree of ventricular enlargement), transducers with different focal depths were employed (55 mm and 65 mm focal depth transducers) as determined by measuring, on a T1-weighted MRI, the distance from the thalamic target to the exterior portion of the temporal bone at its thinnest position. Sonication was administered in 20 blocks of 30 s, with 30 s rest intervals in between, for a total of 10 minutes of (non-continuous) cumulative sonication. Each period of sonication was delivered at a pulse repetition frequency of 100 Hz, a pulse width of 0.5 ms, and thus a 5% duty cycle. This parametrization, which matches that used in prior work,^30,35–37^ has been shown to shorten time-to-awakening following general anesthesia in preclinical models^28^ and is believed to be suppressive of cortical excitability^27,51^ as reflected by the modulation of extracellular GABA concentrations.^26^ With a derated spatial peak temporal average intensity (I_SPTA_) of 14.4 W/cm^2^ and a derated spatial peak pulse average intensity (I_SPPA_) of 0.72 W/cm^2^, the applied sonication falls within the FDA safety envelope.^52^

Prior to sonication, the transducer was coupled to the subject’s head using BrainSonix transmit acoustic standoff pads and a layer of ultrasound gel (Aquasonic 100, Parker Laboratories) to couple the gel pad to the transducer and to the head. While hair and hair follicles are considered to be transparent to acoustic energy,^53,54^ to minimize the adverse effects on acoustic transmission of bubbles forming in the gel between the transducer and the participant’s head,^55^ hair were shaved in correspondence of the transducer position prior to sonication.

Targeting and real-time targeting monitoring was achieved using a Brainsight neuronavigation system (Rogue Research, Inc., Canada) custom fit for use with the BX Pulsar 1002. Optical trackers were placed on a custom transducer holder and on the participant’s head, which was co-registered with the patients T1-weighted image so to allow real-time visualization of the transducer’s position relative to the participant’s brain and the thalamic target.

### Outcome measures

#### Neurobehavioral assessment

The primary measure of interest was the change in maximum CRS-R total score and subscales^40^ after tFUS, as compared to baseline. The CRS-R has been shown to be sensitive to changes in DOC patients following behavioral,^56^ pharmacological,^10,57,58^ and device-based^14–18^ neuromodulatory interventions. While it is well known to be less sensitive than the CRS-R at the lower end of disability, we also acquired the Cisability Rating Scale (DRS)^59^ at enrollment, one week, and one month post-discharge.

#### Electroencephalography (EEG)

EEG is a well-known biomarker of impairment in DOC, with more severe impairments associated with slower oscillatory dynamics.^42,60–63^ Furthermore, given prior data showing an association between the proportion of slow (δ) to fast (β) power-spectrum density (PSD) and the degree of observed thalamic atrophy,^42^ the change in δ/β ratio following tFUS, compared to baseline, was selected as the EEG measure of interest.

#### Positron Emission Tomography

^18^F-FDG PET is a well-known biomarker of impairment in DOC^46,47^ is sensitive to interventions.^11,57^ Consistent with prior work,^46,47^ the primary outcome measure was the change in standard uptake values (SUV) in thalamus and cortex.

#### Polysomnography

Change in sleep stage percentage (N1, N2, N3, REM) after tFUS, compared to baseline, was selected as the variable of interest, as suggested by thalamic DBS work in DOC.^13^

### Data acquisition and analysis

#### Neurobehavioral

Maximum CRS-R total scores in the pre-tFUS and post-tFUS periods were compared using a paired-samples student t-test. Subscale testing was conducted with a Wilcoxon signed-rank test due to violation of normality (as measured by the Shapiro-Wilk test), and corrected for multiplicity with the FDR procedure (at q=0.05).^64^

#### Electrophysiology

EEG data were acquired with a BrainVision actiCHamp (Brain Products) system with 64 Ag/Ag/Cl active electrodes. Before each recording, impedances were confirmed to be below 100 kΩ. Eye-blink and motion artifacts were rejected manually and with an ICA-artifact rejection algorithm. Spectral measures in each frequency band (i.e., δ: 1–4□Hz; θ: 4–8□Hz; α: 8–12□Hz; β: 12–30□Hz) was computed using a Fast Fourier Transformation with the Welch method with a periodogram of 512□ms with 400 ms overlap.^62^ Finally, values were aggregated over epochs using the trimmed mean 80% (mean of the distribution after trimming the 10% lowest and 10% highest values, a robust estimator of central tendency) to obtain a two-dimensional topographical representation over all electrodes per each subject. Data were processed in EEGLAB. To compare the global δ/β ratio in PSD immediately before to immediately after tFUS, a Wilcoxon signed-rank test was used due to violation of normality (as measured by the Shapiro-Wilk test). In addition, the same analysis was also conducted separating the δ/β PSD for frontal, central, temporal and parietal channels (adjusted for multiplicity with an FDR approach).

#### FDG-PET

Brain metabolism data were acquired on a PET/CT system (Biograph 40 TruePoint, Siemens) at the Pomona Valley Hospital Medical Center (PVHMC). Patients were fasted for at least 4 hours and, 60 minutes prior to data acquisition, received a bolus of 3MBq/kg of ^18^F-FDG, administered by the PVHMC radiology personnel following the medical center’s standard operating procedures. Between tracer administration and data acquisition, the patient layed supine and awake (eyes open), in a dimmed, quiet environment. 3D PET data acquisition (at a resolution of 1.34×1.34×1.5 mm) lasted 10 minutes and included a low-dose CT for attenuation correction.^65^ A head holder was used to minimize head-motion during acquisition. Following prior work in DOC,^44,66^ reconstructed in vivo radioactivity concentrations were converted to body weight adjusted standard uptake values (SUV)^67^ as follows:

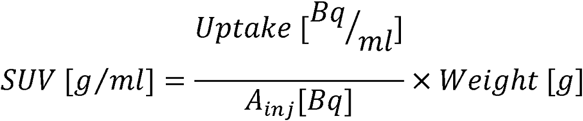

Prior to group analysis, SUV maps were smoothed with a gaussian kernel (5 mm FWHM) and realigned to a cohort-specific PET template (see Supplementary Materials). To compare SUV across the three timepoints (i.e., pre-tFUS; post-tFUS_1_; post-tFUS_2_), data were entered into a group analysis, together with covariates of non-interest (i.e., sex, age, time since injury, etiology [TBI versus NTBI]), using a non-parametric permutation approach as implemented in FSL randomize.^68,69^ Full brain statistics were corrected for multiplicity with a familywise correction at p < 0.05.

#### Polysomnography

On a nightly basis, patients were fit with a Dreem 2 device. This device is a wireless headband worn during sleep which records and analyzes 5 physiological signals *via* three types of sensors: (i) brain cortical activity, with five EEG dry electrodes yielding seven derivations (FpZ-O1, FpZ-O2, FpZ-F7, F8-F7, F7-O1, F8-O2, FpZ-F8; 250 Hz with a 0.4–35 Hz bandpass filter);(ii) movements, position, and breathing frequency via a 3D accelerometer located over the head; and (iii) heart rate, via a red-infrared pulse oximeter located in the frontal band.^48,49^ Prior work has shown that the Dreem2 device is capable of performing high quality automatic sleep staging classification comparable to that obtained with manual expert scoring of medical-grade polysomnography data in healthy volunteers^48^ as well as in clinical populations.^70^ To minimize noise-related variability, for each participant we selected the baseline and pos-tFUS recordings with highest quality index^48^ and at least 120 mins total time.

## Results

### Study population

Sixteen patients (44% Female; Age, M=37.81, SD=15.97) with a chronic DOC (Time Since Injury, M=3.39, SD=1.94 years) secondary to severe brain injury (TBI 44%, non-TBI 56%) were enrolled. The average DRS was 20.44 (SD=3.12, range [17:27]). As shown in the CONSORT diagram (see Supplementary Materials, Figure 2), two patients completed only partially the 10-day protocol. In one case, the family elected to withdraw from the study shortly after undergoing tFUS; in the other case, the patient was withdrawn prior to tFUS due to excessive agitation during the MRI acquisition. The average CRS-R_max_ across the full enrolled population at baseline (N=16) was 10.25 (SD = 2.49).

**Figure 2.**
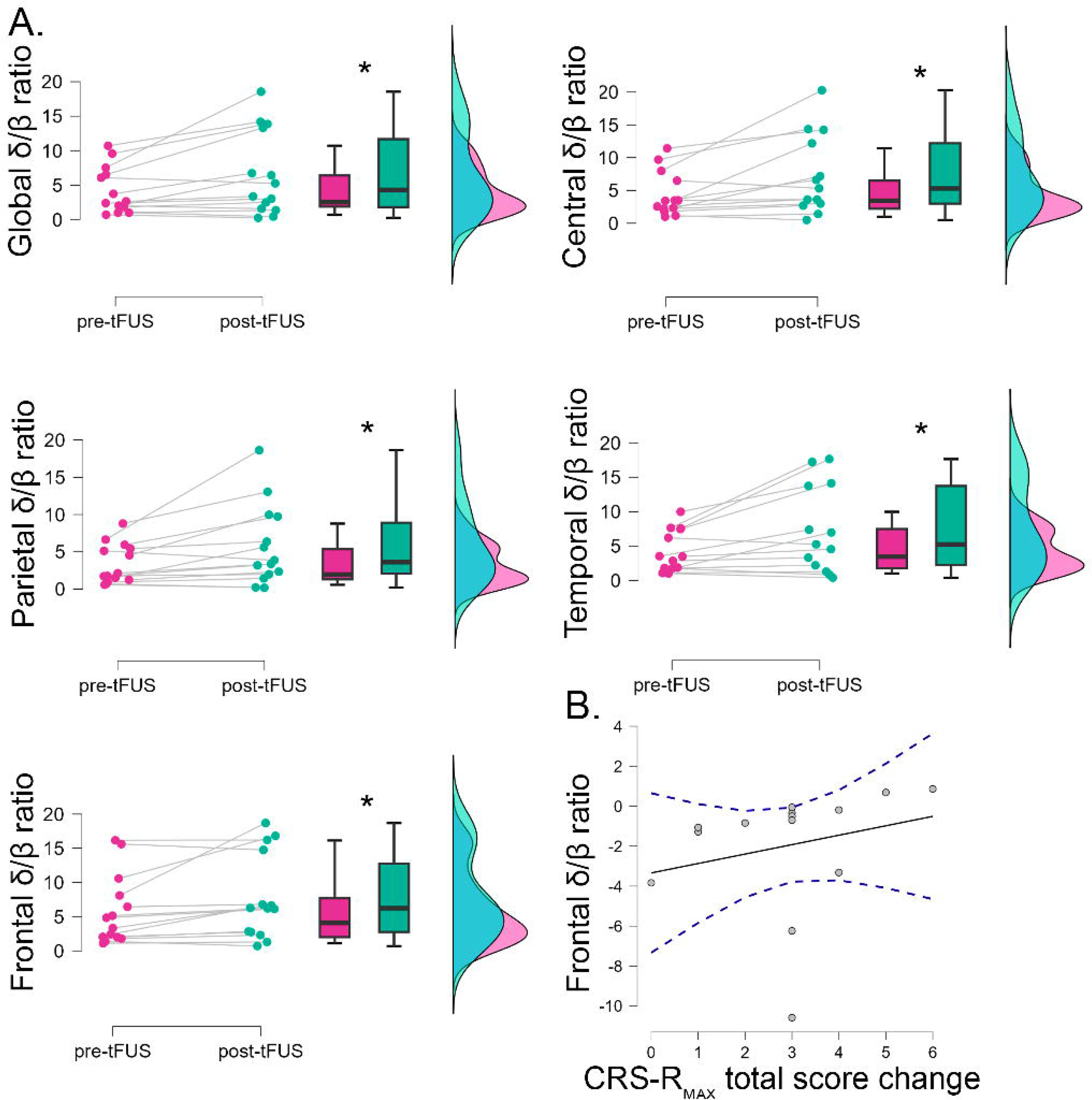
*Electroencephalography results.* (A) Comparison of the power spectrum global, central, parietal, temporal, and frontal δ/β ratio immediately before and immediately after tFUS. (B) Correlation between the magnitude of the observed change in frontal δ/β ratio and the subsequent observed change in maximum CRS-R total score.

### Safety

No severe adverse events were observed. Two adverse events were reported during the 10-day stay (urinary tract infection, hypotension). Independent medical monitor review considered both cases unrelated to the intervention.

### Outcomes

#### Neurobehavioral

The maximum CRS-R total score following tFUS (M=13.27, SD=2.25, range=[9-18]) was significantly greater than at baseline (N=15; M=10.33, SD=2.55; range=[8-16]; t(14)=7.407, p<0.001, Cohen’s d=1.913; see Figure 1A). As depicted in Figure 1B, the change in behavioral responsiveness was first observable within 48 hours post-tFUS and appeared conserved until discharge. Analysis of the individual subscales revealed that visual function following tFUS was significantly increased by a median of 2 points (W=0, z=2.803, p_FDR_=0.018, r_bs_=1; BF=51.78; see Figure 1C). Although a significant increase was also observed for the auditory, communication, and arousal subscales, none of these findings survived correction for multiplicity. Regarding diagnosis, 20% of patients demonstrated improvements (i.e., 3 patients started exhibiting response to command).

#### Electroencephalography

As shown in Figure 2, the global δ/β ratio of the power spectrum increased significantly following tFUS compared to immediately prior to sonication (N=14; Med_change_=0.69; IQR=4.33; W=17, p=0.025, r_rb_=0.68). The significant increase in δ/β ratio following tFUS was observable across frontal (W=13, p_FDR_=0.0173, r_rb_=0.752), central (W=10, p_FDR_=0.0173, r_rb_=0.780), parietal (W=14, p_FDR_=0.0173, r_rb_=0.733), and temporal (W=15, p_FDR_=0.033, r_rb_=0.670) channels, after FDR correction for multiple comparisons. To assess whether the pre-/post-tFUS change in δ/β ratio was associated with changes in neurobehavioral testing, we explored the correlation between the two measures. The increase in δ/β ratio in frontal electrodes was found to be significantly positively associated with the observed change in maximum CRS-R, albeit only marginally after FDR correction for multiplicity (Kendall τ_b_=0.51, p=0.016, p_FDR_=0.060). (See Supplementary Materials for additional EEG analyses.)

#### Metabolism

As shown in Figure 3, glucose metabolism (SUV) was decreased, as compared to baseline, at both post-tFUS PET acquisition. Comparison of each post-tFUS timepoint to baseline resulted in largely overlapping areas of SUV decrease in left and right thalamus, and medial and lateral frontal, parietal, and temporal cortices (see ***Figure 3***). No significant difference was observed when comparing the two post-tFUS datasets, nor when assessing the association between pre-/post-tFUS metabolism change and change in maximum CRS-R total score.

**Figure 3.**
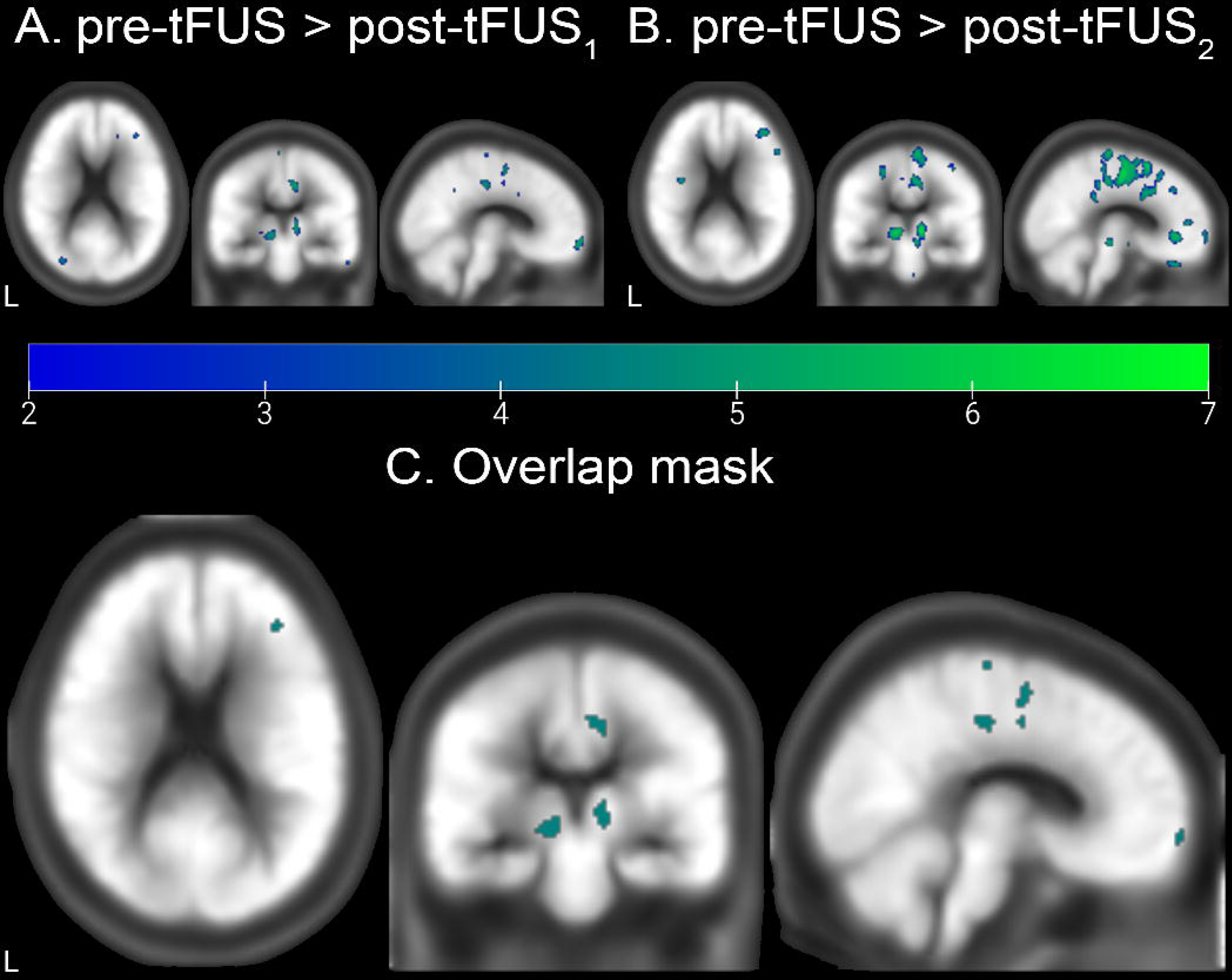
*PET results.* (A) Regions with significant decrease in glucose metabolism (FDG-PET) at the first post-tFUS PET session (post-tFUS_1_) compared to pre-tFUS baseline. (B) Regions with significant decrease in glucose metabolism (FDG-PET) at the second post-tFUS PET session (post-tFUS_2_) compared to pre-tFUS baseline. (C) Conjunction binary mask showing regions exhibiting a significant decrease in glucose metabolism in both contrasts. (As discussed in the main text, no significant difference was detected across the two post-tFUS timepoints.)

#### Polysomnography

Finally, we assessed whether tFUS modified the percentage of sleep spent in each stage (N1, N2, N3, REM). The percentage of time spent in N2 sleep was increased by 33% following tFUS (N=11; t(10)=2.386, p=0.038, Cohen’s d=0.72; BF_10_=2.099; see ***Figure 4***), as compared to baseline. However, this result was not significant after adjusting for multiplicity with FDR-correction. No other association, or correlation with change in behavioral scores, was detected for any of the remaining sleep stages.

**Figure 4.**
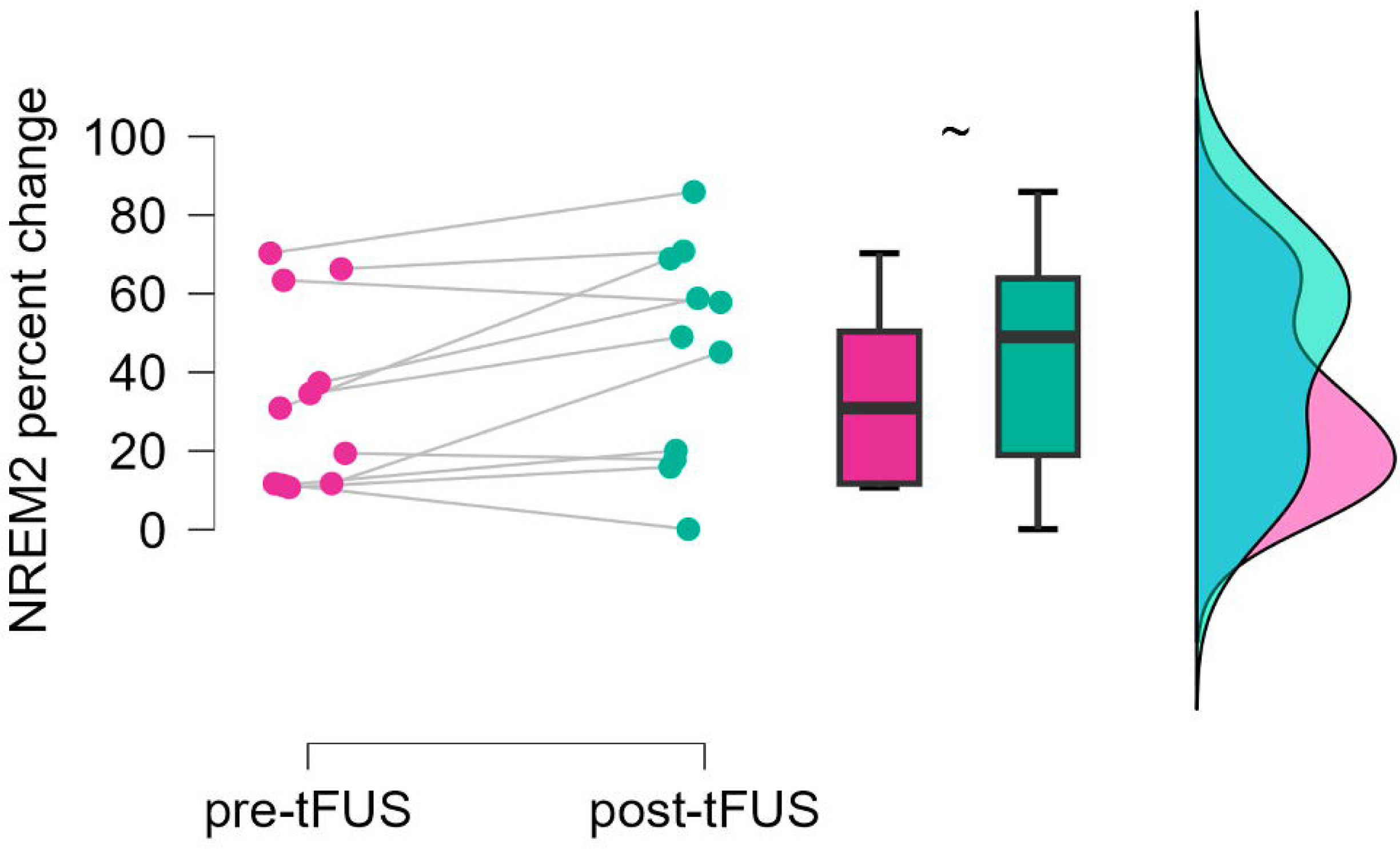
*Polysomnography.* Change in percentage of N2 sleep following tFUS (post-tFUS) compared to baseline (pre-tFUS). (“∼” indicates the difference is significant, but only marginally so after FDR correction for multiplicity.)

#### Long term follow-up

The DRS collected at 1 week and 1 month post tFUS was not significantly different from the DRS at enrollment (F(2,18)=0.982, p>0.05), although the DRS over time did exhibit a significant interaction with sex (F(2,18)=5.726, p=0.012) and a three-way-interaction with sex and etiology (TBI vs NTBI; F(2,18)=5.060, p=0.018).

## Discussion

In this longitudinal, open-label trial, we report the safety and bio-behavioral effects of thalamic low-intensity focused ultrasound in a cohort of patients with chronic DOC. We report three main findings. First, the intervention was safe and feasible; no adverse events related to the sonication were observed, and vital parameters remained stable throughout the protocol, in line with published work.^38^ Second, consistent with our proof-of-concept reports, we observed a significant increase in clinical measures of responsiveness in the week following tFUS,^35–37^ particularly within the visual subscale of the CRS-R. Third, and most notably, we find thalamic tFUS to be associated with a slowing of the scalp EEG signal and a protracted decrease in glucose metabolism in bilateral thalamus and several medial frontal and parietal cortices, among others. Crucially, the magnitude of the EEG slowing in frontal electrodes was positively associated with subsequent behavioral improvement, linking the intervention to the behavioral outcome.

On the one hand, these results are consistent with the preclinical data demonstrating that thalamic sonication at this parametrization (i.e., 100 Hz, 5% duty cycle) enhances recovery of behavioral responsiveness^28^ while suppressing cortical activity.^22,26,27,51^ On the other hand, these findings are counter-intuitive as compared to the excitatory valence of established interventions such as DBS.^14–16^ In this respect, our findings are better aligned with the idea that “silencing” the brain can be beneficial in DOC patients.^71^ While this remains speculative, it is possible that the inhibitory nature of the 5% duty cycle protocol may functionally “reset” thalamocortical activity, by dampening excessive inhibition or by restoring the balance of excitation and inhibition. In turn, this might allow for the re-emergence of the necessary functional substrate for complex behavior to arise—perhaps as a device-based analogue to the paradoxical awakening observed following administration of CNS depressants (e.g., zolpidem).^11,12^

These findings must be interpreted considering certain limitations. As a single-arm trial, we cannot rule out spontaneous recovery or placebo effects, although the chronic nature of our sample (TSI M=3.32 years) and the temporal correlation between physiological changes and behavioral gains mitigate this risk. In addition, open-label studies are known to overestimate effect sizes,^72^ which stresses the need for a double-blind investigation of tFUS. Another limitation is the sample size, which prevents us from formally testing differences in our etiologically heterogeneous sample. Similarly, to enroll 16 patients, we had to evaluate several more, with 24 not meeting inclusion criteria and 10 refusing participation, which adds a level of biased sampling. Lastly, we only tested one tFUS parametrization, and one exposure. Whether other sonication settings or repeated exposures could enhance the effectiveness of tFUS in DOC, remains to be investigated.

In conclusion, this study provides mechanistic evidence that thalamic tFUS can modulate well-validated biomarkers of DOC. The association between thalamic inhibition and behavioral gains complements the prevailing approach to neuromodulation in DOC, suggesting that in chronic patients, dampening thalamic activity may be a path to restoring large-scale network function. These results provide the necessary foundation for a rigorous multicenter, double-blind, randomized clinical trial to validate tFUS as a therapeutic intervention for DOC.

## Data availability statement

De-identified tabulated data can be requested with the authors and shared, upon reasonable request, under an institutional material transfer agreement with Casa Colina Hospital and Centers for Healthcare.

