## Supplementary Materials for "Thalamic sonication in chronic disorders of consciousness: a mechanistic single-arm clinical trial"

### eFigure 1. Schedule of Activity

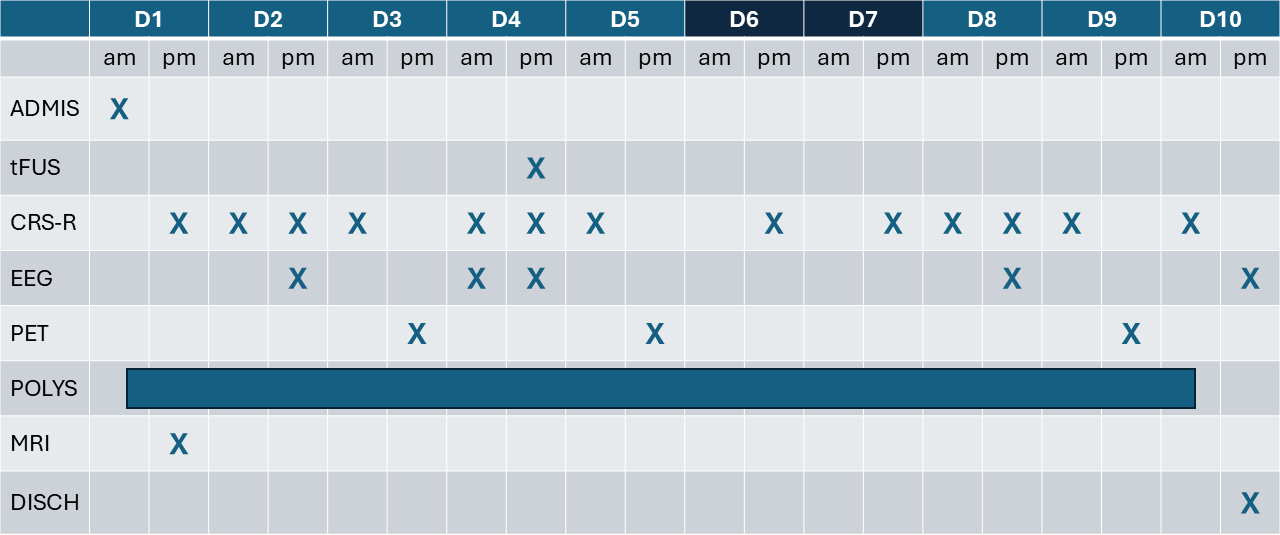

Schedule of activities. (Abbreviations: ADMIS., admission; tFUS, focused ultrasound administration; CRS-R coma recovery scale-revised; EEG, electroencephalography; PET, positron emission tomography; POLYS, polysomnography; MRI, magnetic resonance imaging; DISCH, discharge.)

### eFigure 2. CONSORT diagram

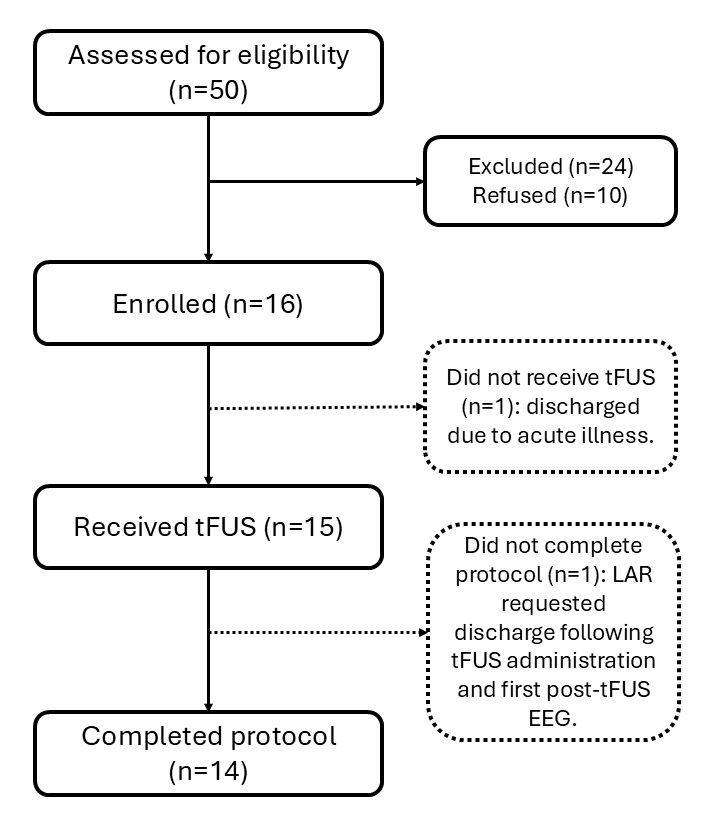

### Cohort-specific template for FDG-PET analysis

A cohort-specific FDG-PET template was constructed from 14 subjects with variable session numbers per subject (range: 1-3 sessions, total: 40 scans). Template creation followed a two-stage procedure to account for within-subject variability and between-subject anatomical differences.

**Within-Subject Registration**. For subjects with multiple sessions, images were rigidly registered (6 degrees of freedom) to the first session using ANTs (Advanced Normalization Tools, v2.x) SyNQuick registration.^1^ This approach corrects for differences in positioning across sessions (which are held on different days) while preserving individual brain morphology. Session-aligned images were then averaged to create a single representative image per subject.

**Template Construction**. The cohort-specific template was created using antsMultivariateTemplateConstruction^2^ with the following parameters: 4 iterations, SyN diffeomorphic registration,^3^ cross-correlation similarity metric (4mm radius), and multi-resolution optimization (4 levels: 6×4×2×1 shrink factors, 2×1×0.5×0 smoothing). To ensure optimal coverage and minimize boundary artifacts, one subject (tFUS008) was selected as the initialization template, based on visual image quality assessment. Prior to template creation, all images were smoothed (2mm FWHM Gaussian kernel) and intensity-normalized to a global mean of 1000 to account for inter-session variability in tracer uptake and scanner calibration.

**Registration to Template Space**. Individual subject means were registered to the final template using SyN diffeomorphic registration. The resulting transformations were then applied to all individual sessions, thus bringing all subject-specific SUV data into a common stereotaxic space that is adapted for this cohort. Final images were intensity-normalized using global mean scaling.

**Quality Control**. Registration quality was assessed through visual inspection and quantitative metrics. Pearson correlation coefficients between each subject's warped image and the final template ranged from 0.91 to 0.97 (mean ± SD: 0.95 ± 0.02), indicating successful spatial normalization. The coefficient of variation map across subjects showed values below 20% in cortical regions, confirming good template stability.

All image processing was performed using ANTs (version 2.), FSL (FMRIB Software Library, version 6.0.4), ^4^ and FreeSurfer (version 7.4).^5^

### Additional EEG analysis

To analyze the full set of EEG recordings spanning all 5 acquisitions, we ran a 4 (regions) × 5 (timepoints) repeated measures ANOVA with sex and etiology (TBI, non TBI) as between subject factors and age and time since injury as additional covariates. The analysis returned a significant effect of time by age (F(4,60)=4.398, p=0.01), time by etiology (F(4,60)=2.866, p=0.05), and the three-way interaction of region, time, and sex (F(12,60)=2.402, p=0.013). The time by age interaction was likely driven by a positive significant correlation between age and δ/β ratio at the fourth timepoint (i.e., post-tFUS2) for central, parietal, and temporal electrodes (r=0.635, p=0.015; r=0.571, p=0.033; r=0.541, p=0.046; respectively). A simple main effects analysis suggests that the significant interaction of time by etiology is due to a significant effect of time for NTBI etiologies (F(4,60)=3.511, p=0.025) and a non-significant simple effect of time for TBI etiologies (F(4,60)=0.052, p>0.05; see Figure 1).

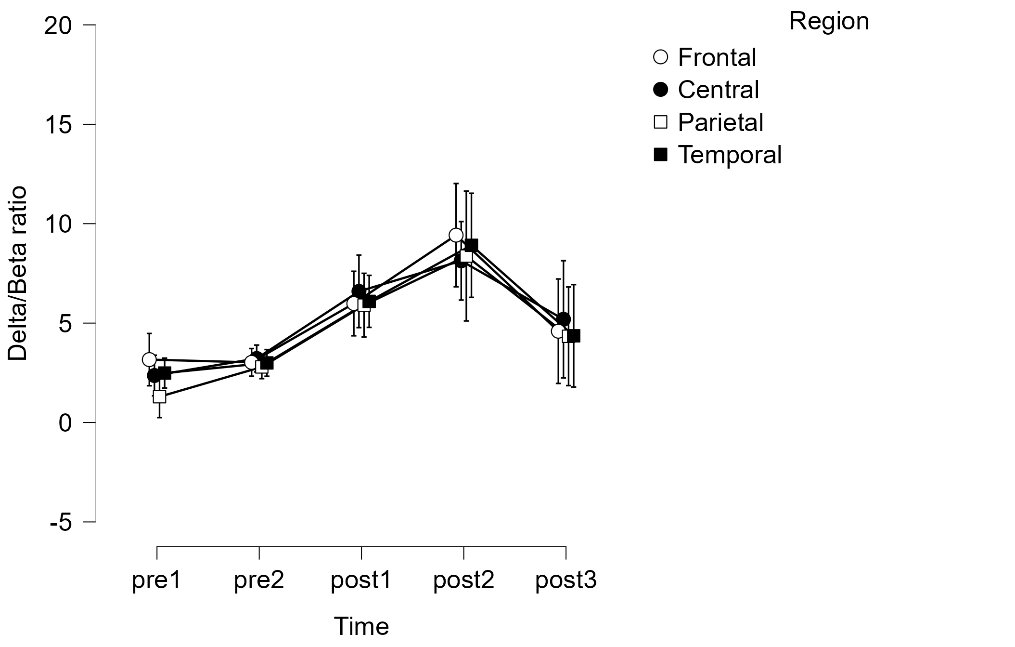

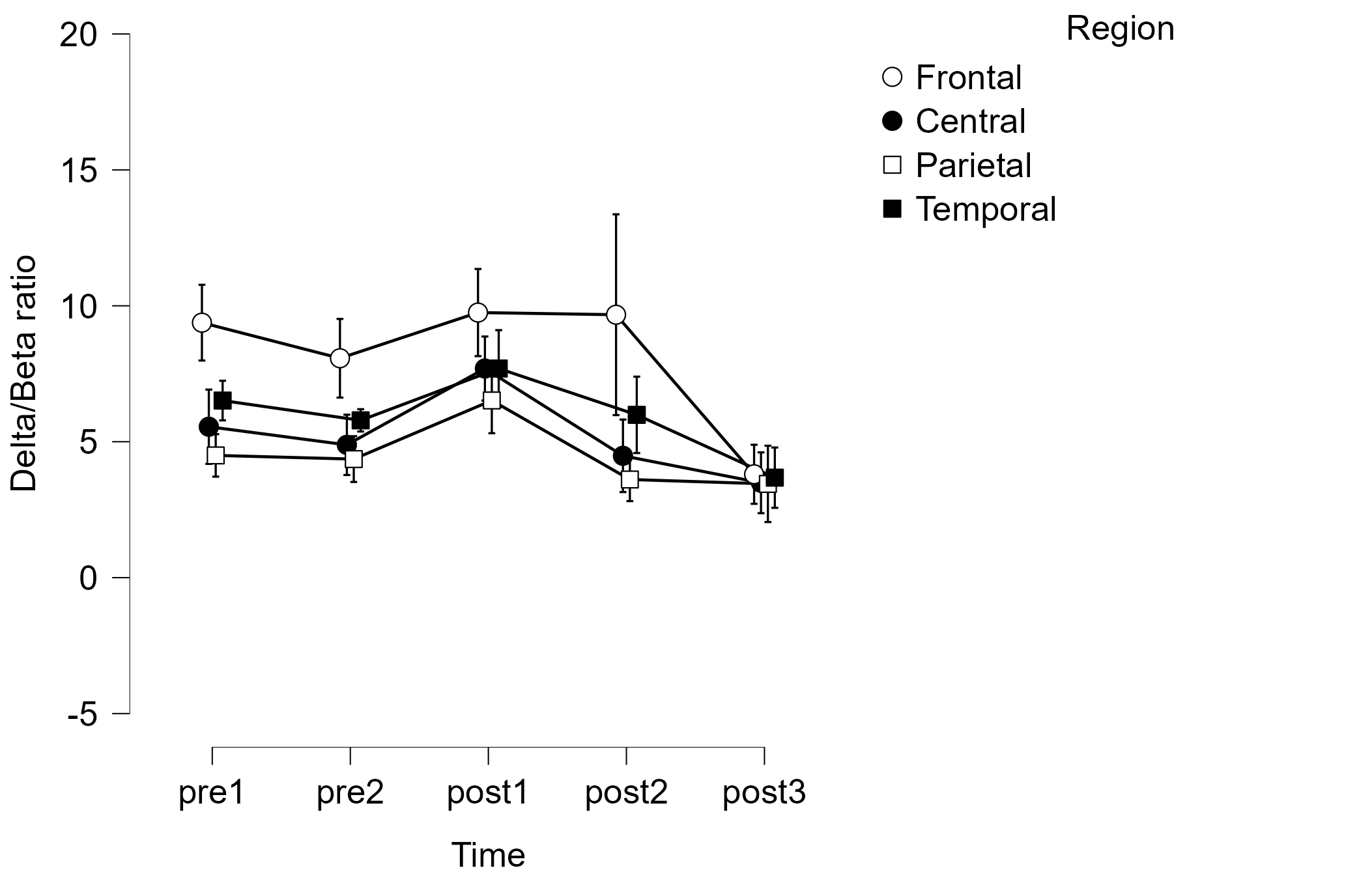

NTBI

TBI

Figure 1. δ/β ratio. Simple main effect of time by etiology.
